# Effectiveness of Ivermectin-Based Multidrug Therapy in Severe Hypoxic Ambulatory COVID-19 Patients

**DOI:** 10.1101/2021.07.06.21259924

**Authors:** Sabine Hazan, Sonya Dave, Anoja W. Gunaratne, Sibasish Dolai, Robert L Clancy, Peter A. McCullough, Thomas J. Borody

**Affiliations:** ProgenaBiome, LLC, 1845 Knoll Dr., Ventura, CA, USA, 93003; Microbiome Research, 1845 Knoll Dr., Ventura, CA, USA, 93003; Centre for Digestive Diseases, 229 Great North Road, Five Dock, NSW, Australia, 2046; Texas A & M College of Medicine, Baylor Dallas Campus, Dallas TX, USA 75226

**Author notes:** Authors’ contributions: SH contributed to and led all aspects of the study, including idea formulation, clinical treatment, study design, Covidex design, analysis, and writing. S Dave carried out data analysis, Covidex design and validation, obtaining control, and contributed to writing. AWG, RLC, and PAM contributed to writing. S Dolai contributed to writing and study design. TJB led all aspects of the study, including idea formulation, study design and writing. Funding: This study was funded by ProgenaBiome, LLC. The authors state that they have obtained appropriate institutional review board approval and have followed principles of the Declaration of Helsinki for all human experimental investigations. Informed consent has been obtained from the participants involved. Data-sharing: The datasets used and/or analysed during the current study are available from the corresponding author on reasonable request.

**Keywords:** SARS-CoV-2, COVID, Coronavirus, Ivermectin, Doxycycline, Zinc

## Abstract

Ivermectin is a safe, inexpensive and effective early COVID-19 treatment validated in 20+ RCTs. Having developed combination therapies for *Helicobacter pylori*, we tested various COVID-19 combinations and describe the most effective. In 24 consecutive COVID-19 subjects with high risk features, hypoxia and untreated moderate-severe symptoms averaging 9 days, we trialed this novel combination comprising ivermectin, doxycycline, zinc, and Vitamins D and C. It was highly effective. All subjects resolved symptoms in 11 days on average, and oxygen saturation improved in 24hrs (87.4% to 93.1%, *p*=0.001). Hospitalizations and deaths were significantly fewer (*p*<0.002 or 0.05, respectively) than in background-matched controls from the CDC database. Triple combination therapy is safe and effective even in moderate-severe patients with hypoxia treated in the outpatient setting.

**Trial Registration:** N/A, see methods.

## Introduction

There is currently a lack of effective treatments for early or ambulatory patients with COVID-19. Patients testing positive are sent home to isolate and generally, no specific line of treatment is prescribed in this phase of the illness. However, there is growing evidence that certain repurposed drugs with good safety profiles, taken early, can significantly improve outcomes and even avoid or delay the need for immune-modulators, antiplatelet / antithrombotic therapy, and the administration of oxygen [1].

Among the most extensively studied of such COVID-19 therapeutics is ivermectin (IVM), a drug that has been used safely in 3.7 billion doses worldwide since 1987 [2-4]. Recently, Dr. Satoshi Omura, the 2015 Nobel prize co-laureate for the discovery of IVM, and colleagues comprehensively reviewed studies to date on IVM activity against COVID-19, concluding that the preponderance of the evidence demonstrated such efficacy [2]. IVM has been tested in more than 20 randomized controlled trials (RCTs) for COVID-19 treatment, with statistically highly significant clinical benefits in almost all of these and a pooled mortality reduction of 78% for the treatment vs. control groups [5]. Five such studies for IVM treatment of COVID-19 recently published in top-tier medical journals have all shown multiple clinical benefits for IVM vs. controls, most of these with high statistical significance on the order of *p* < 0.002 [6-10]. IVM is well tolerated at much greater than the standard single dose of 200 μg/kg [11,12] and has been used in RCTs for COVID-19 treatment at cumulative doses of 1,500 μg/kg [13], 1,600 μg/kg [14], and 3,000 μg/kg [15] over 4 or 5 days either without or with mild and transient adverse effects. Not surprisingly, IVM has become extensively used in the prevention and early disease management of COVID-19, particularly in non-Western countries.

Despite this strong evidence of clinical benefit in COVID-19 for IVM therapy, variation in therapeutic regimens especially with respect to addition of a broad spectrum antibiotic and zinc, has led to confusion as to how best to manage acute infections. Indeed, the most impressive of the early ambulatory multi-drug therapy, claiming 87% and 75% reductions in hospitalization and deaths, respectively, both with a P value of 0.001, in 869 high risk subjects, left optimal management strategy unclear due to a mixed use of IVM and hydroxychloroquine (HCQ) [16]. There is an immediate need for an effective, safe, and practically available combination therapy formulated on the basis of the best available data.

At a cellular level, IVM modulates communication between the cytoplasm and nucleus, creating a hostile environment for assemblage of virus, while at the same time reducing cytokine-mediated inflammation. In addition, IVM inhibits pathology following infection with the COVID-19 virus, by specifically blocking binding of the virus “Spike” protein to the ACE2 receptor. Finally, IVM has been associated with favorable changes in cellular innate immunity [17].

Our group has been developing antiviral drug combinations for COVID-19 and found IVM to be particularly effective as a co-therapy for use early in COVID-19 to shorten the time to symptom resolution and to prevent hospitalization. IVM used alone can at times be only partially effective but not curative [6,1,19] yet a higher dose of IVM plus azithromycin and zinc has achieved a 92% mortality reduction vs. controls [14]. Thus, we chose a combination of safe and widely available medications, approved for other indications and without drug-drug interactions or QT prolongation that inhibits intracellular virus replication and possess some anti-inflammatory properties.

The use of combination therapy for intracellular bacterial infections is not new and has been used successfully to treat Tuberculosis, *Helicobacter pylori* infections, leprosy and intracellular viral infections such as *Hepatitis B & C -*where a single component of the combination therapies is rarely curative. In some viral infections e.g. HIV, even combined multiple antiviral drugs cannot completely cure but suppress the viral load perpetually. IVM is best known for its broad-spectrum efficacy for parasite infections, its high cure rate and limited drug resistance when used in combination [20]. Although useful, IVM used alone is not the ‘magic bullet’. Combinations can help lower individual doses and reduce side effects. To cover all age group requirements, we combined IVM with doxycycline and zinc as active components and with Vitamins D and C as replacement ‘excipients’ given to supplement common clinical deficiencies in the aged.

This study reports the use of the above combination therapy in consecutive, ambulatory, complex, at times profoundly hypoxic patients whose oxygen saturation (SpO_2_) was as low as 73%. Participants were treated by an experienced clinical trials team within Ventura Clinical Trials Inc..

## Methods

### Subjects

Subjects were identified from patients referred by physicians, or word-of-mouth in Los Angeles, Ventura County, CA, and other states in the USA. These patients were referred to participate in clinical trials under clinicaltrials.gov ID NCT04482686 (which is a double-blind Randomized Control Trial). However some did not qualify for this trial as their oxygen saturation was less than 90%, and were deemed too sick to enter a placebo-controlled trial. Given they were excluded and refused to go to the hospital, they were treated off-label via telemedicine, during Aug. 2020 and Feb. 2021. Subjects were given the opportunity to participate in this open label trial with IRB oversight once the diagnosis was made via swab RT-qPCR testing once the diagnosis was made via swab RT-qPCR testing. Inclusion criteria were as follows 1) positive PCR for COVID-19; 2) informed consent; 3) age > 18 years, and 4) agreement to practice two highly effective methods of birth control if of childbearing potential. Exclusion criteria included 1) allergies or drug interactions with the combination therapy components; 2) listed comorbidities, including seizure risk; and 3) pregnancy.

### Treatment

Treatment began as soon as practical, within 72 hrs. of patients presenting to Ventura Clinical Trials. All screened subjects met the inclusion criteria and were enrolled consecutively. Treatment was defined as ‘IVM Combination Therapy’ (ICT) and consisted of ten days of oral: Doxycycline (100mg twice a day), IVM (12mg on day 1, day 4, and day 8), Zinc (25mg twice a day), Vitamin D_3_ (1500 IU twice a day) and Vitamin C (1500mg twice a day). ICT was given daily for ten days only.

Two patients (#10 and #23) received an initial treatment on day one of 36 mg IVM (rather than 12mg) due to particularly low SpO_2_ or expected clinical need.

### Monitoring

Subjects self-recorded symptoms in their daily logs (supplementary 1) for the first 10 days. Electrocardiograms (EKGs), blood pressure, temperature (reported in ^0^F) and SpO_2_, were collected via provided medical equipment at home. On days 1, 5, 10 and 30, SARS-CoV-2 testing swabs were self-collected by subjects and sent to pathology for testing. Pregnancy tests were performed as appropriate.

### Endpoints

Endpoints were 1) time from presentation to negative SARS-CoV-2 PCR; 2) time from presentation to symptom resolution; 3) progression to hospitalization; 4) patient survival.

### Externally Controlled Trial (ECT) Arm

Given the challenges for COVID-19 of enrolling high risk severely hypoxic patients in an open-label trial with an untreated control arm, our treated group arm survival was compared to the control group survival rate in the general population. This ECT, also known as a synthetic control arm, was calculated from the public CDC database of COVID-19 subjects [21]. Available information includes age range, presence of any chronic condition (COVID-19-vulnerability or otherwise, conditions not specified), date of infection, and whether the COVID-19 diagnosis was laboratory-confirmed. We used information from all subjects who met the following criteria: 1) age 50+ years; 2) laboratory-confirmed COVID-19 diagnosis; 3) death/survival, race, and sex status available and known; 4) infection prior to March 2021; and 5) subject had any co-morbidities. This synthetic control arm development was carried out after our clinical data were obtained, and so selection criteria chose the control subjects closely matched to the subjects in our study, all of whom had some underlying condition and a large majority were over 50 years of age. The CDC database was analysed using CSViewer vs. 1.3 (EasyMorph Inc, Toronto, ON, CA, http://easymorph.com).

### Covidex calculations and statistics

Covidex and Covidex-F are ambulatory SARS-CoV-2 infection disease severity measures that we developed and validated in this study. They are weighted particularly to emphasize SpO_2_, and Covidex-F includes a variable for body temperature.

Covidex score = 1 pt. (if history of sleep apnea) + 1 pt. (if history of COPD) + 1 pt. (if history of cardiovascular disease) + 1pt. (if history of asthma) + 1pt (if history of prior clots, ischemia or stroke) + 1pt (if obese, i.e. 30 kg/m^2^ < BMI < 40 kg/m^2^) + 2pts (if severely obese, i.e. BMI > 40) + 1pt (if age > 60 years) + [95-(SpO_2_ as a percentage)]pts. For instance, a hypothetical patient with a history of asthma and morbid obesity with a SpO_2_prior to treatment of 85% would have a Covidex score of 1 (for asthma) + 2 (for obesity) + 10 (for SpO_2_of 85%) = 12 pts.

Covidex-F score = Covidex score + 1 pt. (if temperature on presentation between 99.5^0^F and 100.4 ^0^F) + 2 pts. (if temp on presentation between 100.4^0^F and 103.5^0^F) + 3 pts. (if temp on presentation > 103.5^0^F).

Best-fit lines were made to assess the correlation between Covidex scores and time from treatment to symptom resolution. Regression was carried out in Prism version 8 (GraphPad Prism software for Windows, San Diego, California USA, www.graphpad.com) using least square regression without weighting or special handling of outliers. All graphs were prepared by and statistical analysis done using GraphPad Prism version 8, and the error bar are indicative of the standard error of the mean (SEM).

## Results

Table 1 lists all subjects in the study, two of which did not consent to ICT treatment (subjects #10 and #26), and their associated race, gender, symptoms, fever, and other clinical notes. All subjects had COVID-19-related symptoms on presentation, and the symptom range was broad with several showing shortness of breath (SOB). The vast majority of subjects, 21 of 24 (87.50%), had fever on presentation with a mean temperature for all 24 subjects of 101.2 + 0.32 ^0^F. Specifically, 1/24 (4.17%) had low grade fever (99.5-100.4), 18/24 (75.00 %) had medium grade fever (100.5-103.4), and 2/24 (8.33%) had high grade fever (> 103.4) ^0^F.

**Table 1:** Listing of subjects and COVID-associated symptoms on presentation and other characteristics. O_2_% Sat = SpO_2_ just before time of treatment initiation. O_2_% Sat post 24 hrs. = SpO_2_ 24 hrs. after treatment initiation. Rx Start date = Day 1 of ICT administration. SOB = Shortness of Breath

| ID | Age | Race | Gender | Symptoms | Temp.<br>( ° F) | O2 %<br>sat. | O2 % sat.<br>post 24<br>hrs. | Rx.<br>start<br>date | Symptom<br>resolution<br>date | PCR pos.<br>date | PCR neg.<br>date |
| --- | --- | --- | --- | --- | --- | --- | --- | --- | --- | --- | --- |
| 1 | 65-69 | caucasian | M | runny nose, sore throat, dizzy, low energy | 99.3 | 90 | 94 | 12/nm/2020 | 12/nm/2020 | 11/nm/2020 | 1/nm/2021 |
| 2 | 60-64 | caucasian | M | SOB, chest congestion, productive cough, nausea, vomit | 105 | 77 | 87 | 12/nm/2020 | 12/nm/2020 | 11/nm/2020 | 12/nm/2020 |
| 3 | 75-79 | caucasian | M | low energy | 101 | 88 | 96 | 10/nm/2020 | 11/nm/2020 | 10/nm/2020 | 10/nm/2020 |
| 4 | 65-69 | caucasian | F | loss of appetite, cough, chills, SOB | 101 | 97 | 96 | 10/nm/2020 | 10/nm/2020 | 10/nm/2020 | 10/nm/2020 |
| 5 | 65-69 | caucasian | F | vomit, weak, body aches, anosmia | 101 | 89 | 95 | 12/nm/2020 | 12/nm/2020 | 12/nm/2020 | n/a |
| 6 | 40-45 | caucasian | F | PE, headache, body ache, cough | 101 | 88 | 94 | 1/nm/2021 | 2/nm/2021 | 1/nm/2021 | 2/nm/2021 |
| 7 | 60-64 | caucasian | M | productive cough, headache | 102 | 86.5 | 91 | 11/nm/2020 | 12/nm/2020 | 11/nm/2020 | 12/nm/2020 |
| 8 | 55-59 | caucasian | M | cough, nasal congestion, SOB, body ache | 102 | 88 | 96 | 10/nm/2020 | 11/nm/2020 | 10/nm/2020 | 11/nm/2020 |
| 9 | 90-94 | hispanic | F | low energy, SOB, confusion, loss of appetite, shaking | 102 | 88 | 94 | 1/nm/2021 | 1/nm/2021 | 12/nm/2020 | n/a |
| 10 | 65-69 | hispanic | M | cough, SOB, respiratory failure | 100.6 | 72 | 87 | declined | death | 12/nm/2020 | n/a |
| 11 | 60-64 | hispanic | F | cough, SOB | 102 | 90 | 96 | 1/nm/2021 | 1/nm/2021 | 12/nm/2020 | n/a |
| 12 | 45-49 | hispanic | M | SOB | 104 | 84 | 91 | 12/nm/2020 | 12/nm/2020 | 12/nm/2020 | n/a |
| 13 | 65-69 | caucasian | F | cough, congestion, rash | 102 | 88 | 91 | 11/nm/2020 | 12/nm/2020 | 11/nm/2020 | n/a |
| 14 | 65-69 | caucasian | M | post-nasal drip, cough, sinus pain | 98 | 88 | 91 | 11/nm/2020 | 12/nm/2020 | 11/nm/2020 | n/a |
| 15 | 70-74 | hispanic | M | low energy, productive cough, anosmia | 101 | 88 | n/a | 12/nm/2020 | 1/nm/2021 | 12/nm/2020 | n/a |
| 16 | 65-69 | hispanic | F | dry cough, body aches, low energy, anosmia | 100 | 88 | n/a | 12/nm/2020 | 1/nm/2021 | 12/nm/2020 | n/a |
| 17 | 45-49 | caucasian | F | diarrhea, rash, renal pain | 102 | 87 | 94 | 8/nm/2020 | 8/nm/2020 | 7/nm/2020 | n/a |
| 18 | 85-89 | caucasian | M | cough, fever, low energy | 102 | 88 | 95 | 1/nm/2021 | 1/nm/2021 | 1/nm/2021 | 1/nm/2021 |
| 19 | 55-59 | caucasian | F | stomach pain, diarrhea, cough, rash | 102 | 90 | 95 | 9/nm/2020 | 9/nm/2020 | 8/nm/2020 | n/a |
| 20 | 50-54 | muslim | M | cough, fever, loss of appetite, chills | 101.2 | 88 | n/a | 10/nm/2020 | 10/nm/2020 | 10/nm/2020 | 10/nm/2020 |
| 21 | 90-94 | caucasian | M | low energy | 102 | 85 | 91 | 2/nm/2021 | 2/nm/2021 | 2/nm/2021 | 2/nm/2021 |
| 22 | 60-64 | hispanic | M | cough, low energy, loss of appetite | 101.3 | 90 | 96 | 2/nm/2021 | 2/nm/2021 | 2/nm/2021 | n/a |
| 23 | 55-59 | hispanic | M | cough, SOB | 98 | 73 | 90 | 1/nm/2021 | 2/nm/2021 | 2/nm/2021 | 1/nm/2021 |
| 24 | 45-49 | hispanic | F | chest pain, SOB | 98.6 | 90 | n/a | 2/nm/2021 | 2/nm/2021 | 2/nm/2021 | n/a |
| 25 | 85-89 | hispanic | M | severe SOB, low energy, trouble walking | 101.6 | 90 | 95 | 2/nm/2021 | 3/nm/2021 | 2/nm/2021 | n/a |
| 26 | 85-89 | caucasian | M | SOB | 102 | 88 | n/a | declined | death | 10/nm/2020 | n/a |

Table 2 summarizes the demographics and past medical history (PMH) of subjects who consented to treatment (total n=24, 2 additional subjects who declined treatment are excluded). Notably, patients were older (a known COVID-19 vulnerability) with a mean age of 66 + 2.75 years, and a range of 43 to 94 years (Table 2A). The population of the 24 subjects consenting to treatment (not subjects #10 and #26, Table 1) was 63% males. Death of untreated subjects #10 and #26 excluded downstream analysis in the other figures.

**Table 2.**
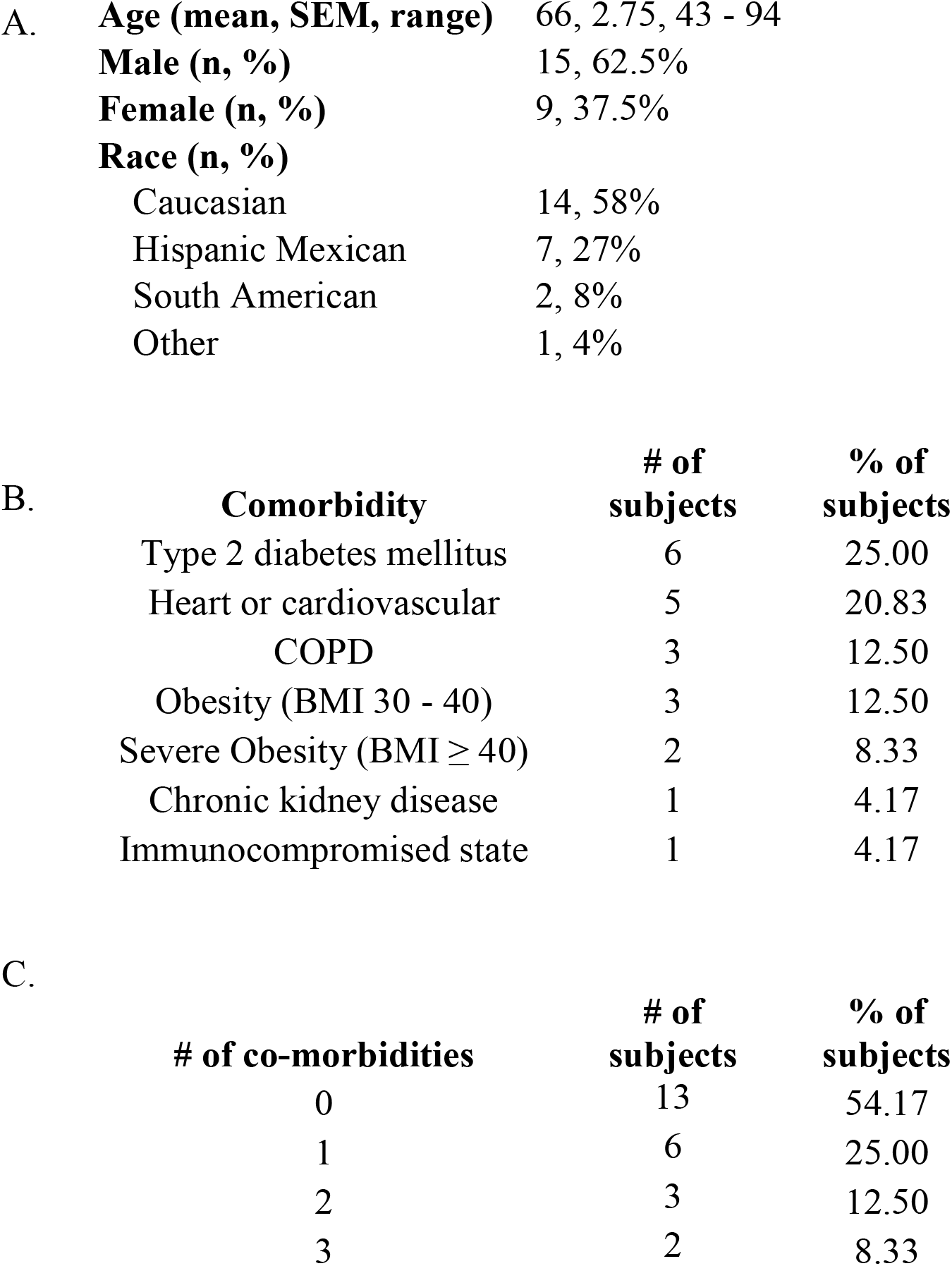
A. Age and demographics of subjects. B. Prevalence of COVID susceptible co-morbidities. C. Number of concurrent comorbidities in subjects.

Table 2B lists the number of patients who had comorbidities associated with COVID-19 vulnerability, based on CDC guidelines[22]. These comorbidities are chronic kidney disease, chronic obstructive pulmonary disease (COPD), Down syndrome, cardiovascular disease, immunocompromised state including HIV, obesity (body mass index [BMI] of 30 kg/m^2^ or higher but < 40 kg/m^2^), severe obesity (BMI ≥ 40 kg/m^2^), pregnancy, sickle cell disease, smoking, type 2 diabetes. Of note, no subjects had cancer, Down syndrome, or sickle cell disease and none were pregnant nor were smokers.

Many subjects had multiple comorbidities associated with COVID-19 vulnerability, as outlined in Table 2B. In total, 11/24 (45.83%) subjects had COVID-19-vulnerable comorbidities of which 3 (12.50%) had 2 separate comorbidities, and 2 (8.33%) had 3 co-morbidities.

A minority of subjects (n = 7) had other COVID-19 treatment(s) prior to and/or during ICT administration, namely Remdesivir (n = 1 subject), involvement in a placebo-controlled trials of HAZDpaC (n = 4 subjects, trial clinicaltrials.gov NCT04334512; may have been given treatment or placebo) and hydroxychloroquine (HCQ, n = 3 subjects).

Fig. 1 demonstrates all subjects recovered from COVID-19, typically within one to two weeks. Fig. 1A shows various durations for each subject and average values (one outlier excluded). Time from onset of symptoms to treatment initiation is shown in column 1 and averages 9.2 + 2.1 days. Time from start of treatment to symptom resolution was11.6 + 1.4 days. Time from first positive to first negative PCR was 16.9 + 1.6 days and is less than three weeks. Time from start of treatment to first negative PCR was 11.5 + 1.6 days and is also less than three weeks.

**Figure 1:**
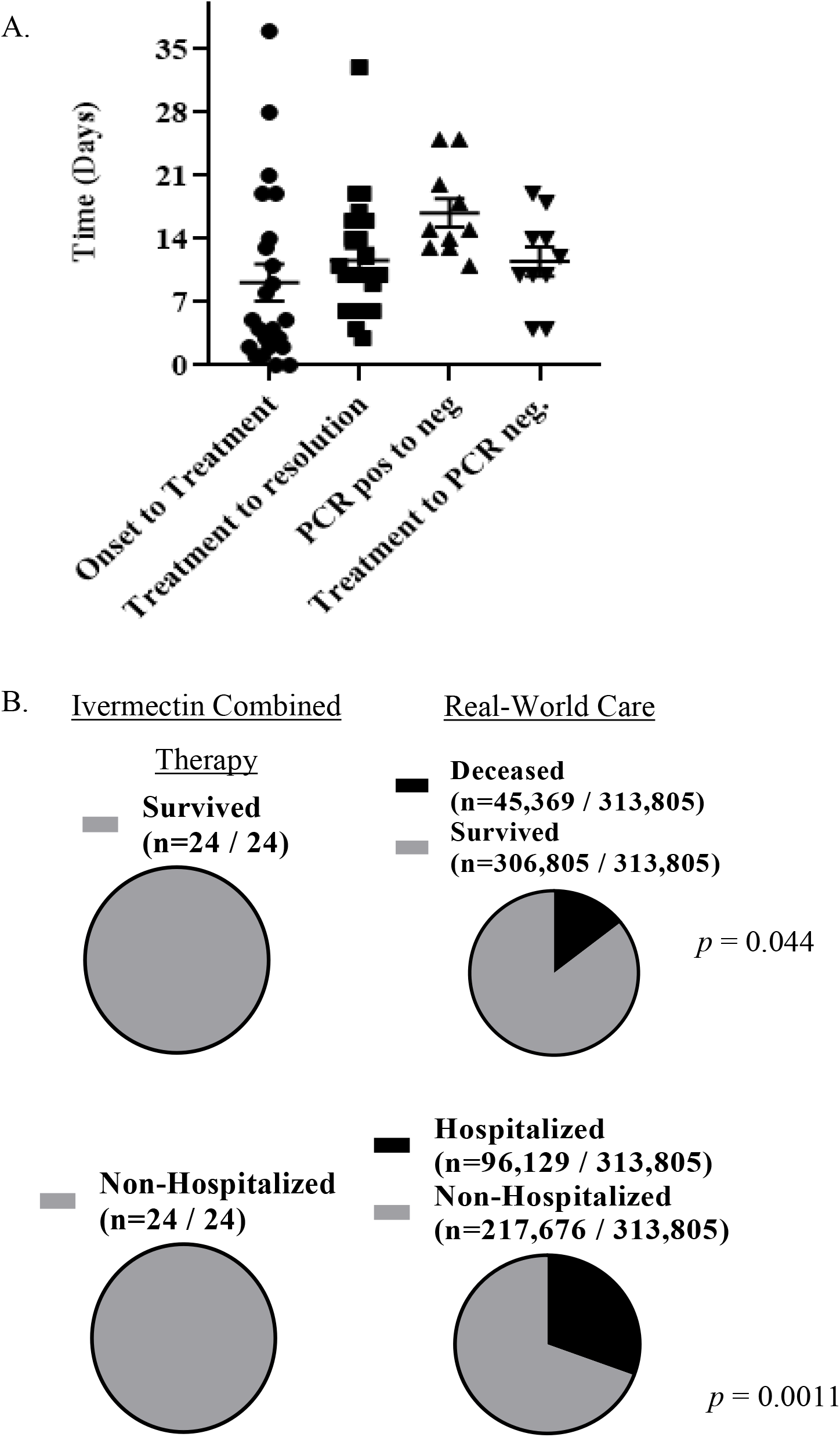
Complete recovery was seen in all patients within 1 to 3 weeks. A. Time in days to various stages of symptom onset and resolution. Nearly all subjects resolved symptoms and became PCR negative in 3 weeks Col 1: Symptom onset to start of treatment (n = 23, mean = 9.17 + 2.05) Col 2: Start of treatment to resolution of symptoms. (n = 23, mean = 11.61 + 1.38) Col 3: First PCR positive to first PCR negative (n = 10, mean = 16.90 + 1.58) Col 4: Start of treatment to first PCR negative (n =10, mean = 11.50 + 1.60) B. Top, 100% survival rate was seen in patients, which is significantly higher (*p* = 0.044 via χ^2^ test) than synthetic control from CDC database of equivalent or less COVID-vulnerable subjects. Bottom, No (0%) patients required hospitalization, which is significantly less (*p =* 0.0011 via χ^2^ test) than synthetic control from database.

Fig. 1B shows that 100% of subjects survived COVID-19, without need for hospitalization or ventilator use. As noted in Table 2, many of these subjects were older and with comorbidities. When compared to the synthetic control arm, derived from the CDC database (see methods, and as follows), this was a significant increase in survival rate (*p* = 0.044) and decrease in hospitalization rate *(p* = 0.0011), evaluated via χ^2^ test. Of note, the patients in this CDC database likely did receive treatment, of an unknown nature. Thus, the survival rate of this synthetic control reflects the “typical” survival rate in the USA, which is significantly less than the 100% survival rate observed on ICT.

The 100% survival rate on ICT was compared with survival rates from the CDC database of COVID-19 subjects. 356,424 control subjects were obtained, based on qualification criteria described in the methods section. These criteria focused on older subjects (50+ years), similar to our study population, that also had underlying conditions. One should note that the underlying condition criteria information available in this database refers to chronic conditions of any type, whether or not the condition induces COVID-19-vulnerability. With this definition, 100% of control subjects and ICT-treated subjects had underlying conditions of any type.

Our critical finding in Fig. 2 (see also Table 2), showed that 23/24 patients were hypoxic with SpO_2_ < 90%. Some subjects consenting to treatment had SpO_2_ as low as 73%, 77%, 84% and 85% on presentation. As a whole, shown in Fig. 2A, the SpO_2_ of subjects was significantly less than 95%, the defined point of cure (95% CI of mean SpO_2_= 85.5 to 89.4, Mean = 87.4 + 0.93%). Subjects’ SpO_2_ increased within 24 hrs. of treatment. Their mean SpO_2_ before treatment (for subjects with data before and after 24 hrs.) was 86.5% *+* 1.3, and after 24 hrs. of treatment, 93.1% + 0.63, a highly significant and rapid increase (p <0.001). SpO_2_ then continued to rise. Treatment continued for 10 days reaching the point of successful treatment or cure, which was SpO_2_ > 95%. Successful treatment was reached for all subjects. That is, there was a 100% restoration rate in terms of SpO_2_. No patient who accepted treatment required hospitalization.

**Figure 2.**
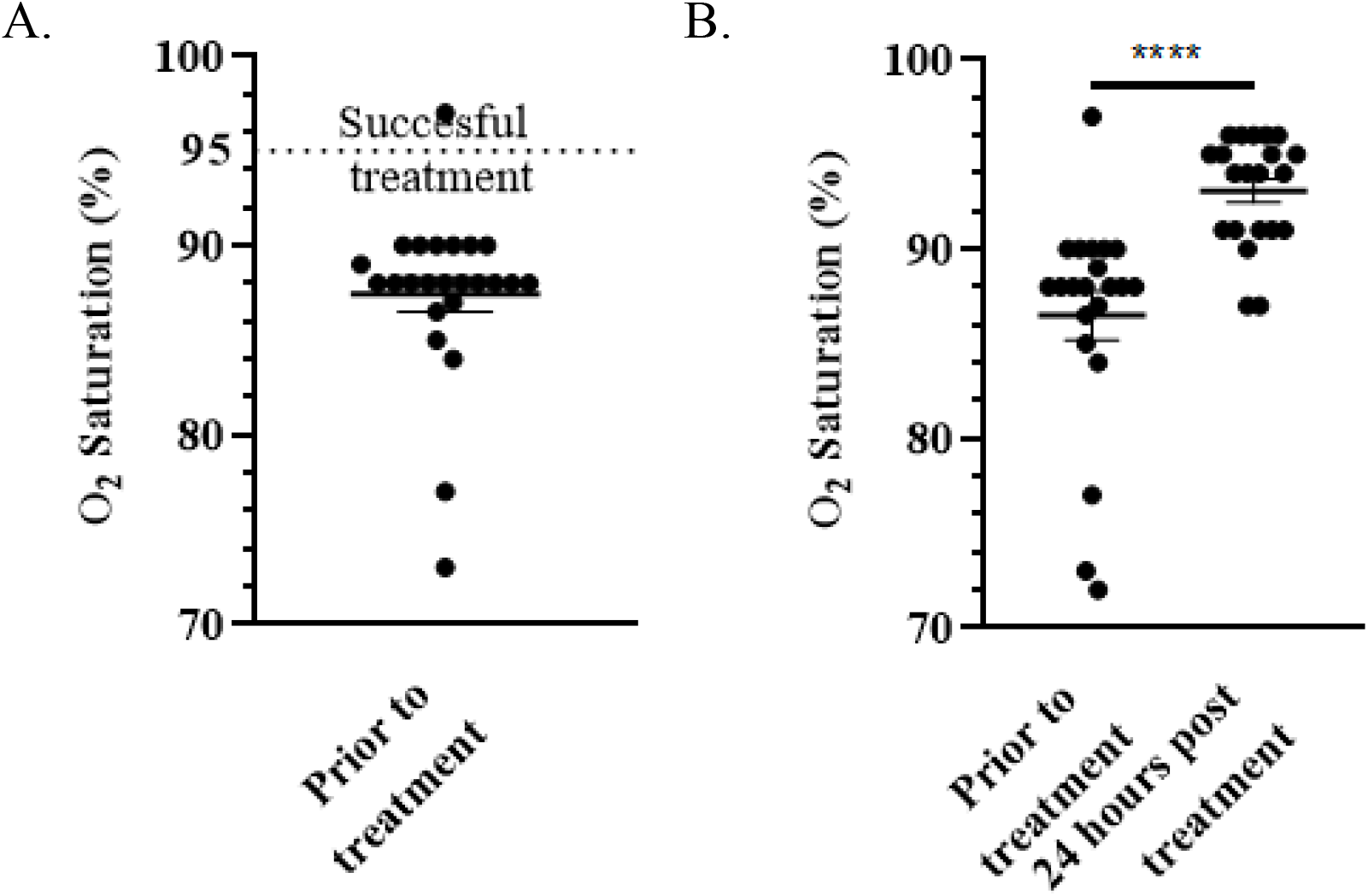
A. O_2_ saturation prior to treatment was significantly (*p* < 0.05) less than 95%, the defined successful treatment reached by all subjects. B. O_2_ saturation significantly increased in subjects 24 hours after treatment (paired *t*-test, *p* < 0.001, only subjects with data before and after treatment included). O_2_ saturation continued to rise, and until the defined cure of greater than 95% O_2_ saturation.

Two subjects (#10 and #26) declined treatment. These subjects did not recover SpO_2_ and died from COVID-19 infection. An adverse drug event of dizziness was reported by one subject, who nevertheless continued with treatment successfully otherwise.

Fig. 3 shows the validation of the Covidex and Covidex-F scores we developed, defined in our methods. These scores provide an index of COVID-19 predicted severity, based on past medical history, O_2_ just before treatment and fever grade on presentation. The mean Covidex score was 10.34 + 1.08 and the mean Covidex-F score was 11.63 + 1.13, with 87% and 85% of the score points, respectively, coming from the contribution of the SpO_2_ term. In other words, approximately 80 to 90 % of this score is weighted towards SpO_2_. It should also be noted, the PMH aspects that contribute to this score emphasize respiratory, cardiovascular, and obesity histories and differ from CDC defined COVID-19-vulnerabilities.

**Figure 3:**
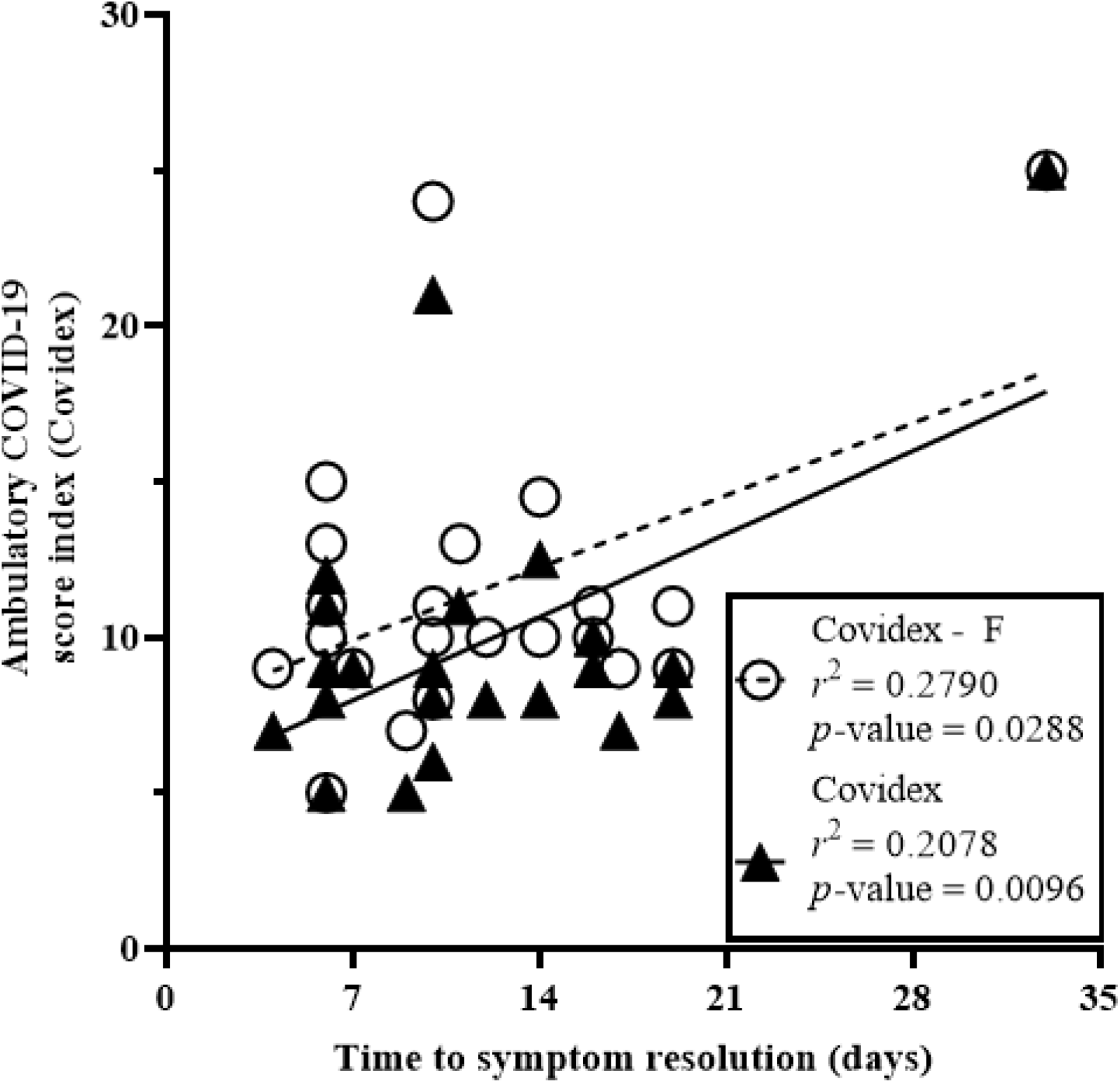
Ambulatory COVID scores, Covidex and Covidex-F (see methods for definition; Covidex-F includes fever measure) vs. time from start of treatment to symptom resolution. There was significant relation between either Covidex score (Covidex, *p* = 0.0288; Covidex-F, *p* = 0.0096) and treatment resolution time.

Covidex and Covidex-F are both plotted vs. time from treatment to symptom resolution. Both shows statistically significant correlations (Covidex: *p* = 0.0096, *r*^2^ = 0.2078; Covidex-F: *p* = 0.0288, *r*^2^ = 0.2790), indicating that either Covidex or Covidex-F is associated with and may predict time to symptom resolution.

## Discussion

We report for the first time a highly effective ICT which led to 100% survival and cure in unselected ambulatory ‘moderate to severely’ ill COVID-19 patients with hypoxia managed as outpatients. Given our experience developing combination therapies for *H. pylori*, we trialed a number of different IVM-based combinations on ambulatory COVID-19 patients, searching for a cure, and found the above combination to be the best as a ‘foundation therapy’ for COVID 19. Understanding that a personalized approach may require added HCQ or other components in some patients much like *H*.*pylori* resistant to triple therapy sometimes requires quadruple therapy. Hypoxia is a demonstrated predictor of COVID-19 mortality [23]. For example, several of these patients had profound hypoxia, measured by oximetry, at 73%, 77%, 84% and 85% on presentation. Despite a symptom to treatment delay of 9.2 days, our treatment brought rapid improvement – beginning in some within 12 hrs. with a mean SpO_2_ rising from 86.5 to 93.1 in the first 24 hrs. There was a parallel improvement in the symptoms including loss of cough, fever and tiredness. Also, the time from the start of treatment to the first negative PCR averaged 11.5 + 1.6 days. Generally, such ill patients would have been admitted to the hospital, yet all those treated with the ICT avoided hospitalization and none died.

Turning to the ECT ‘synthetic control arm’ it is clear that the ICT was statistically superior to the control arm even though a small patient group was reported. The very low adverse effects from reported studies and this treatment group supports the use of ICT if clinical symptoms and risk factors for COVID-19 progression are present, even in cases with PCR pending results. ECT arms are now increasingly used, especially where the control arm or ‘standard-of-care’ arm may have a fatal outcome[24].

The rationale for using combination antiviral therapy is based on our growing understanding that intracellular infections – bacterial or viral, cannot be reliably cured using a single drug. It is also based on our knowledge that IVM resistance is common. Hence, there is no single ‘silver bullet’ for COVID-19, and the indiscriminate use of IVM *alone* could induce COVID-19 resistance by generating drug-resistant strains. Resistance to IVM was the case when used alone in scabies, nematodes, strongyloidiasis, microfilaridermias, onchocerca and volvulus [19,25-31]. We are now seeing resistance develop even in combined therapies using two antibiotics for *H. pylori*. This teaches us to use IVM in COVID-19 only in combination therapies, especially so, with growing reports of mutant strains resulting in vaccine breakthrough infections [32]. Hence, the proposed combination therapy was developed to induce *cure* more rapidly, *prevent* resistance, and *overcome mutant strain* emergence -no replication no mutation.

Regarding strategies in the development of combination therapies, intracellular coronavirus replication requires several active drugs to inhibit viral replication. IVM, doxycycline and zinc all individually inhibit coronavirus replication and, although there are other candidates, we have proposed the above combination based on efficacy, component safety profiles, inexpensive nature, and lack of drug-drug interaction. The combination of IVM and doxycycline has also been demonstrated to act in synergy against COVID-19 [33]. This combination also appears to overcome the need for high doses of IVM identified by Caly and colleagues when used alone[34]. Further, given that zinc plays a key role in antiviral activity [34536] it would combine well with the ionophores (IVM and doxycycline) to increase its intracellular concentration and expedite viral clearance [37]. We have also assessed drug-drug interactions and found that the combination of zinc with IVM and doxycycline has no reported interactions. Additionally, each of these drugs has a low adverse side effects profile and no QT prolongation as reported with azithromycin.

Overall, based on the current literature, a 10-day combination therapy of IVM, doxycycline and zinc will not only improve symptoms [6,7] but also accelerate recovery from COVID-19. We have chosen a safe IVM dosage approved for parasites of 36mg over 10 days, and this dose has been shown to be both effective and safe in COVID-19 treatments [38]. The staggered IVM dosage over 10 days is proposed based on the half-life clearance of the drug in plasma (up to 66 hrs.)[39]. The proposed duration would allow constant availability of adequate plasma level IVM to facilitate zinc entry into the cells. Hence, the above rationale explains why some publications have already shown that IVM alone is not adequate to cure COVID-19 [6,18,19] while a multidrug regimen is likely to be more efficacious [40].

While underpowered, this study enrolled *consecutive subjects* into study and did not bias subject selection from different time points. Many of the enrolled subjects were profoundly ill with subjective assessments that may have resulted in hospital admission and/or intubation. Yet, ICT anti-viral activity appears to have rapidly restored SpO_2_ and reversed other symptoms which could not be explained simply by the developing immunity.

A weakness of this study is the lack of a concomitantly enrolled control arm. However, given the potential fatal outcomes Tess Lawrie et al. indicated [41] *‘it is no longer ethical to approve the use of a control arm as so many profoundly ill patients in the control arm would die’*, as did our two subjects who declined treatment [42]. Hence our study has made use of the ECT or ‘synthetic’ control arm which has enabled us to make matched age and comorbidity comparisons. Institutional Review Boards should now include the provision to include available synthetic arms and reject trials that include in a COVID-19 trial a control arm as published by Lawrie[41].

This study builds on an extensive literature, to provide a practical inexpensive, safe, readily available and highly effective triple therapy aiming to prevent resistance and one that can confidently be used as a routine treatment for outpatient COVID-19.

## Data Availability

The datasets used and/or analysed during the current study are available from the corresponding author on reasonable request.

## Declarations

### Ethics approval and consent to participate

All subjects were explained the study and provided written informed consent. This study was approved by E&I Review Services (https://www.eandireview.com/) as study # 210006.

### Consent for publication

All authors have read and agreed to the final submitted contents of this publication.

## Acknowledgements

The authors would like to thank Margaux Alvaran for technical assistance.

## List of abbreviations

χ^2^: test (Chi-Square Test)
CDC: (Center for Disease Control)
CI: (Confidence Interval)
COVID-19: (Coronavirus, SARS-COV-2)
Hrs.: (Hours)
HAZDpaC: (Hydroxychloroquine,Azithromycin,Vit D, Zinc Pack)
*H. pylori*: (*Helicobacter pylori*)
ICT: (Ivermectin Combination Therapy)
PMH: (Past Medical History)
pt: (point)
pts: (points)
SEM: (Standard Error of Mean)
SOB: (Shortness of breath)
SpO_2_: (Percent Saturation Peripheral Oxygen)
temp: (Body temperature in ^0^F)

## Figure and table legends

**Supplementary Figure 1:**
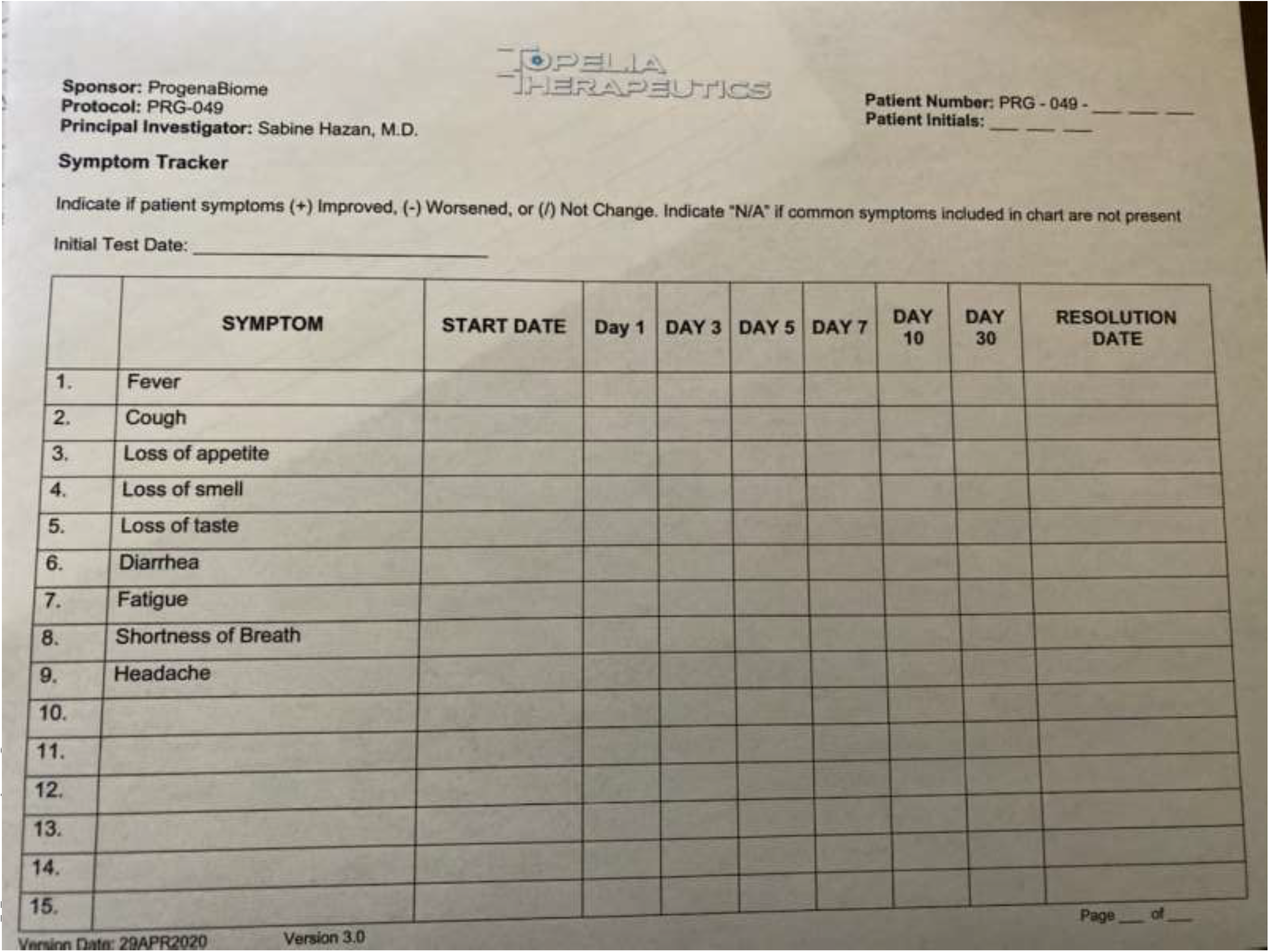
Example of symptom log sheet used by subjects.

